# An ancestry-enriched PIEZO1 missense variant biases HbA1c-based diagnosis of prediabetes and type 2 diabetes in South Asians

**DOI:** 10.64898/2026.03.27.26348321

**Authors:** Miriam Samuel, Daniel Stow, Vi Bui, Margherita Bigossi, Sam Hodgson, Susan Martin, Jana Soenksen, Camila Armirola-Ricaurte, Stuart Rison, Julia Cassasco-Zanini, Benjamin M Jacobs, Genes & Health Research Team, Viswanathan Baskar, Venkatesan Radha, Jebarani Saravanan, Taeko Becque, Viswanathan Mohan, Ranjit Mohan Anjana, David A van Heel, Rohini Mathur, Trevelyan J McKinley, Veline L’Esperance, Moneeza Siddiqui, Inês Barroso, Sarah Finer

## Abstract

**Background:** Glycated haemoglobin (HbA1c) underpins type 2 diabetes (T2D) and prediabetes management worldwide and reflects both glycaemia and erythrocyte biology. A missense variant in *PIEZO1* (rs563555492_T_), carried by 1 in 12 south Asians, has been associated with a nonglycemic reduction in HbA1c. We aimed to further characterise this association and evaluate its clinical consequences.

**Methods:** We undertook genetic and linked health data analyses across two cohorts: 19,898 (37.4% female) South Indians from the Madras Diabetes Research Foundation (MDRF) and 43,011 (54.4% female) British Bangladeshis and British Pakistanis in Genes & Health. In MDRF, we tested associations with glycaemic and erythrocytic traits using additive genetic models. In Genes & Health we modelled diagnosis of prediabetes, T2D, and diabetic eye disease using flexible parametric survival models. Ten-year absolute risks were estimated for a population aged 40-50 years.

**Findings:** *PIEZO1* rs563555492_T_ was associated with erythrocytic traits and lower HbA1c, but not with fasting glucose, postprandial glucose, or C-peptide. This variant reduced risk of prediabetes (HR 0·63, 95% CI 0·58-0·69) and T2D (0·85, 0·78-0·93) diagnosis, and increased risk of diabetic eye disease among individuals with T2D (1·20, 1·01-1·43). Modelling suggested approximately 1,019 missed prediabetes and 303 missed T2D diagnoses per 100,000 adults over 10 years.

**Interpretation:** An ancestry enriched *PIEZO1* variant is associated with lower HbA1c independent of glycaemia, reduced prediabetes and T2D diagnosis suggesting delayed detection, and increased complication risk. Reliance on HbA1c may systematically underestimate glycaemic risk in a substantial minority of south Asians.

**Funding:** The Wellcome Trust; NIHR

**Research in Context:** *Evidence before this study:* We searched PubMed from database inception to Nov 1, 2025, for studies examining PIEZO1, Glycated Haemoglobin (HbA1c) and type 2 diabetes (T2D) related outcomes. Search terms included “PIEZO1”, “haemoly*”, “hemol*”, “erythrocyte*”, “HbA1c”, “glycated haemoglobin”, “glycated hemoglobin”, “type 2 diabetes”, “prediabetes”, “retinopathy”, and related terms. HbA1c is widely used to guide prevention, diagnosis, and management of T2D, but variants affecting erythrocyte structure or lifespan can alter its relationship with glycaemia. A missense variant in *PIEZO1* (rs563555492_T_), carried by one in twelve south Asians, has been associated with lower HbA1c and erythrocytic biomarkers without differences in fasting glucose and with older age at T2D diagnosis, suggesting delayed diagnosis in UK-based cohorts. However, uncertainties remain. Associations have not been replicated outside the UK to confirm generalisability. Experimental animal models suggest PIEZO1 channels may influence pancreatic insulin secretion, but this has not been evaluated in human cohorts. Finally, the potential clinical and health-economic consequences associated with this variant remain largely unexplored.

*Added value of this study:* We replicated associations between rs563555492_T_, lower HbA1c, and erythrocytic changes in a geographically distinct south Asian cohort based in South India. We found no association with fasting glucose, postprandial glucose, or C-peptide concentrations, supporting a predominantly erythrocytic mechanism. We examined the clinical and economic consequences of this variant on south Asians in a large UK population-based cohort, following introduction of HbA1c-based diagnostic criteria. Carriers experienced delayed diagnosis of prediabetes and T2D, and there was some evidence of increased risk of diabetic eye disease. In a modelled population aged 40-50 years, the variant was associated with approximately 1,019 missed prediabetes diagnoses and 303 missed T2D diagnoses per 100,000 adults over 10 years. Because prediabetes diagnosis initiates referral to prevention programmes, missed diagnoses represent lost opportunities for disease prevention. In a UK NHS setting, we estimated the associated opportunity cost to be between £970,000 - £1,390,000 per 100,000 individuals over a 10-year horizon.

*Implications of all the available evidence:* A south Asian ancestry enriched *PIEZO1* variant can disrupt the relationship between HbA1c and glycaemia and delay detection of prediabetes and T2D. Reliance on HbA1c alone may underestimate glycaemic risk in a substantial minority of individuals of south Asian ancestry, with potential implications for prevention strategies, risk of complications and health care costs. Alternative biomarkers of glycaemia or individualisation of HbA1c may help ensure timely and accurate diabetes detection in populations where this variant is common.

## Introduction

Type 2 diabetes (T2D) affects approximately 537 million adults worldwide and is a leading cause of cardiovascular disease, kidney failure, and vision loss.^1^ People of South Asian ancestry experience disproportionately high rates of microvascular and macrovascular complications.^2-4^ Prevention, early detection and management for both prediabetes and T2D is critical to reduce the long-term complications and mitigate substantial health-care costs associated with T2D.^5-7^

Glycated haemoglobin (HbA1c) is internationally recommended for the diagnosis^8,9^ and monitoring^10^ of T2D and for identifying individuals with prediabetes who are eligible for preventative interventions.^6,7,11^ HbA1c is an indirect measure of glucose, reflecting average glycaemia by measuring the proportion of haemoglobin that has undergone glycation. Consequently, non-glycaemic determinants of HbA1c, such as erythrocyte turnover and haemoglobin biology, have the potential to affect the accuracy of HbA1c.^12^

Recent human genetic studies have identified genetic variants in ancestrally diverse populations that are associated with lower HbA1c without corresponding differences in fasting glucose.^13,14^ In UK-based cohorts, a south Asian enriched missense variant in *PIEZO1* (rs563555492_T_), carried by 1 in 12 south Asians, but rare in other ancestral populations,^15^ has been associated with a non-glycaemic reduction in HbA1c, variations in erythrocyte-related biomarkers and older age of T2D diagnosis.^13,16,17^ *PIEZO1* encodes a mechanosensitive cell membrane receptor expressed on erythrocytes and other tissues.^18,19^ Mutations within this gene are known to cause erythrocytic changes and haemolysis.^20,21^

However, uncertainties remain. First, replication of the genetic associations in populations outside the UK is required to assess whether environmental context modifies the effect. Second, animal models suggest pancreatic PIEZO1 receptors directly influence insulin secretion,^22^ but human data are lacking on associations between the effect allele (rs563555492_T_) and direct markers of insulin secretion. Third, associations between the variant and prediabetes diagnosis, a critical window for preventative intervention, has not been evaluated. Finally, it is unknown whether delayed diagnosis associated with this variant translates into increased risk of diabetes-related complications or measurable economic consequences at the population level.

We therefore integrated genomic and phenotypic data from two large south Asian cohorts. We first assessed whether previously reported genetic associations with the PIEZO1 variant could be replicated in an ancestrally and geographically distinct South Indian cohort and leveraged deep glycaemic phenotyping data to evaluate its association with previously uncharacterised glycaemic traits. We then utilised longitudinal health data in a large, longitudinal cohort UK-based south Asians to assess whether the variant was associated delayed diagnosis of prediabetes and T2D, and greater complication risk in individuals with T2D. Finally, we projected the economic impact of delayed prediabetes detection attributable to this variant.

## Methods

We conducted an integrated genetic and health data analysis. Analyses were undertaken in two south Asian cohorts. Participants from the Madras Diabetes Research Foundation (MDRF) in South India contributed to analyses of associations between the rs563555492 variant and glycaemic and erythrocytic biomarkers. British Bangladeshi and British Pakistani participants in Genes & Health (G&H) contributed to analyses of associations with prediabetes and T2D diagnosis, diabetes-related complications, and downstream health economic implications within a health system that predominantly relies on HbA1c for diabetes diagnosis and management.^23^

### Study Cohorts

#### Madras Diabetes Research Foundation (MDRF)

MDRF is the research wing of Dr Mohan’s Diabetes Specialities Centre (DMDSC), a large network of diabetes hospitals and clinics based across South India. Study design, data linkage, and genotyping procedures have been described previously.^24-26^ Ethics approval was granted by the National Institute for Health Research Global Health Research Unit on Global Diabetes Outcomes Research Institutional Ethics Committee of the Madras Diabetes Research Foundation (IRB00002640, granted 24 August 2017). Participants provide informed consent for the use of their health records in research. We included 19,898 individuals with linked genetic and health record data who attended their first diabetes clinic consultation within one year of T2D diagnosis.

#### Genes & Health (G&H)

G&H recruits UK-based volunteers aged ≥16 years, identifying as British Bangladeshi or British Pakistani, and has sites in London, Bradford, and Manchester.^27^ Ethical approval for the study was provided by the South East London National Research Ethics Committee (14/LO/1240). Participants consent to secure, anonymised electronic health record data linkage from multiple healthcare providers and provide a saliva sample for DNA extraction and genomic analyses. We used Whole Exome Sequencing data and longitudinal health data from primary healthcare providers in the December 2024 data release available for 43,811 participants.

### Genetic data

MDRF participants were genotyped using Illumina Global Screening Arrays (GSA) v1.0 or v2.0. We excluded samples with a call rate <95% and genetically inferred sex discordance with phenotype data, batch effects, heterozygosity >3 SDs, and sample duplicates (identity by descent [IBD] score >0.8). We excluded single nucleotide polymorphisms (SNPs) with <97% call rate and Hardy-Weinberg equilibrium p < 1 × 10^−6^ (autosomal variants only). Quality control assessment was performed independently for MDRF cohorts before and after phasing and imputation against the Haplotype Reference Consortium (HRC) version r1.1 panel. The imputation quality for rs563555492_T_ was 97%.

For G&H, high-depth Whole Exome Sequencing was performed using Twist reagents and Illumina 150bp paired-end Novaseq-6000 sequencing at the Broad Institute (USA). Further methods have been described elsewhere.^28^

### Clinical biomarker measurements

In MDRF, routinely collected clinical biomarkers at the time of the first diabetes clinic consultation included HbA1c, fasting and postprandial plasma glucose, fasting and stimulated C-peptide. Biomarker assay procedures have been published previously.^29^ (Anjana, Baskar et al. 2020) Associations between rs563555492_T_ and rank-based inverse normal transformed biochemical data in MDRF was assessed using an additive model adjusted for age, age^2^, sex and five genetically defined principal components. We estimated allelic effects using a linear mixed model as implemented in Regenie v3.4.1,^30^ to account for relatedness and population stratification.

### Clinical outcomes

Clinical outcomes were assessed in G&H using linked health record data. Diagnoses of prediabetes, T2D, and diabetic eye disease were defined using validated code lists (S1 Supplementary). Diabetes-related screening and management in England and Wales are embedded within incentivised national quality improvement frameworks linked to clinical coding, therefore incident events were defined as the first recorded diagnostic code. Follow-up for incident prediabetes and T2D analyses commenced on Jan 1, 2014, corresponding to when WHO guidelines lead to HbA1c being implemented for T2D diagnosis,^8^ and the national rollout of the NHS Health Check, a national screening programme incorporating T2D.^31^ For analyses of diabetic eye disease, individuals with incident T2D diagnosed on or after Jan 1, 2014 were included.

### Survival modelling

Complete case analysis was undertaken. All participants had recorded date of birth. Individuals with genetically ambiguous sex or ancestry were excluded. Participants were censored at death, deregistration, or end of follow up defined as 1^st^ December 2024. Follow-up for incident prediabetes and T2D analyses commenced on Jan 1, 2014, corresponding to adoption of HbA1c-based diagnostic criteria following the 2011 WHO recommendation supporting HbA1c for diabetes diagnosis,^8^ national rollout of the NHS health check, a national screening programme incorporating T2D.^31^ For analyses of diabetic eye disease, individuals with incident T2D diagnosed on or after Jan 1^st^, 2014 were included and follow up commenced on the date of T2D diagnosis.

Time to prediabetes, T2D and diabetic eye disease diagnosis were analysed using flexible parametric survival models^32^ in R v4.4.1. Models were adjusted for age, genetically inferred sex, and principal component-defined ancestry group. Age was modelled using restricted cubic splines to allow for non-linearity using the RMS package. Marginal absolute risk estimates were derived for a modelled population aged 40-50 years at study entry, corresponding to the recommended age range for the first NHS Health Check,^31^ using the standsurv command from the flexsurv package. Further model diagnostics and sensitivity analyses are described in the supplementary material (S3).

### Economic impact modelling

We estimated the projected economic impact of delayed prediabetes detection associated with the rs563555492_T_ allele from the perspective of the UK National Health Service. In England and Wales, a coded diagnosis of prediabetes triggers referral to the NHS Diabetes Prevention Programme, participation in which is embedded within incentivised primary care quality frameworks. Published modelling has estimated that referral of 100,000 individuals to the programme is associated with £135,755 in cost savings and 40·8 quality-adjusted life years (QALYs) gained.^5^ We therefore treated missed prediabetes diagnoses as a foregone benefit (opportunity cost). Using the estimated carrier prevalence and modelled 10-year attributable risk of missed prediabetes diagnosis among individuals aged 40-50 years at study entry, we projected the economic impact of these missed referrals over a 10-year horizon per 100,000 individuals in the UK.

## Results

A total of 63,709 participants with genomic and linked health data contributed to the analyses, including 19,898 from MDRF and 43,811 from G&H. In MDRF, mean age was 49.0 years (SD 10·6), 7,435 (37·4%) were women, and all participants were of Indian ancestry. In G&H, mean age was 40·5 years (SD 13·9), 23,847 (54·4%) were women, and 29,106 (66·4%) were of Bangladeshi and 14,705 (33·6%) of Pakistani PCA-derived ancestry. The rs563555492_T_ allele frequency was 4·1% in G&H and 3·7% in MDRF (Table 1).

**Table 1.** Demographics of the study population with genomic and linked electronic health data.

|  | Madras Diabetes<br>Research Foundation | Genes & Health |  |  |
| --- | --- | --- | --- | --- |
|  |  | All | Bangladeshi | Pakistani |
| Total Population (N) | 19,898 | 43,811 | 29,106 | 14,705 |
| Sex |  |  |  |  |
| Female (N (%)) | 7,435 (37.4) | 23,847 (54.4) | 15,681 (53.9) | 8,166 (55.5) |
| Male (N (%)) | 12,463 (62.6) | 19,964 (45.6) | 13,425 (46.1) | 6,539 (44.5) |
| Age at study entry in years<br>(median (IQR)) | 49.6 (15.3) | 38.8 (17.5) | 38.5 (16.3) | 39.7 (20.5) |
| Effect Allele (rs563555492<sub>T</sub>)<br>Frequency (%) | 3.7 | 4.1 | 4.4 | 3.8 |

We examined the association of rs563555492_T_ with fasting and post-prandial measures of glucose and insulin secretion (C-peptide) using biomarker data in MDRF. In additive genetic models of rank inverse normalised traits, the effect allele was associated with lower HbA1c (n=18,951, β −0·15 SD units, 95% CI −0·20 to −0·10), lower mean corpuscular volume (n=17,238, β −0·22, −0·28 to −0·17) and raised total bilirubin (n=17,770, β 0·12, 0·08 to 0·17) but not with fasting (n=18,956, β 0·03, −0·02 to 0·08) or postprandial glucose (n=18,950, β 0·02, −0·03 to 0·07). No evidence of association was observed for fasting c-peptide (n=14,155, β 0·01, −0·05 to 0·07) or stimulated (post-prandial) c-peptide (n=14,255, β −0·01, −0·06 to 0·05).

**Figure 1.**
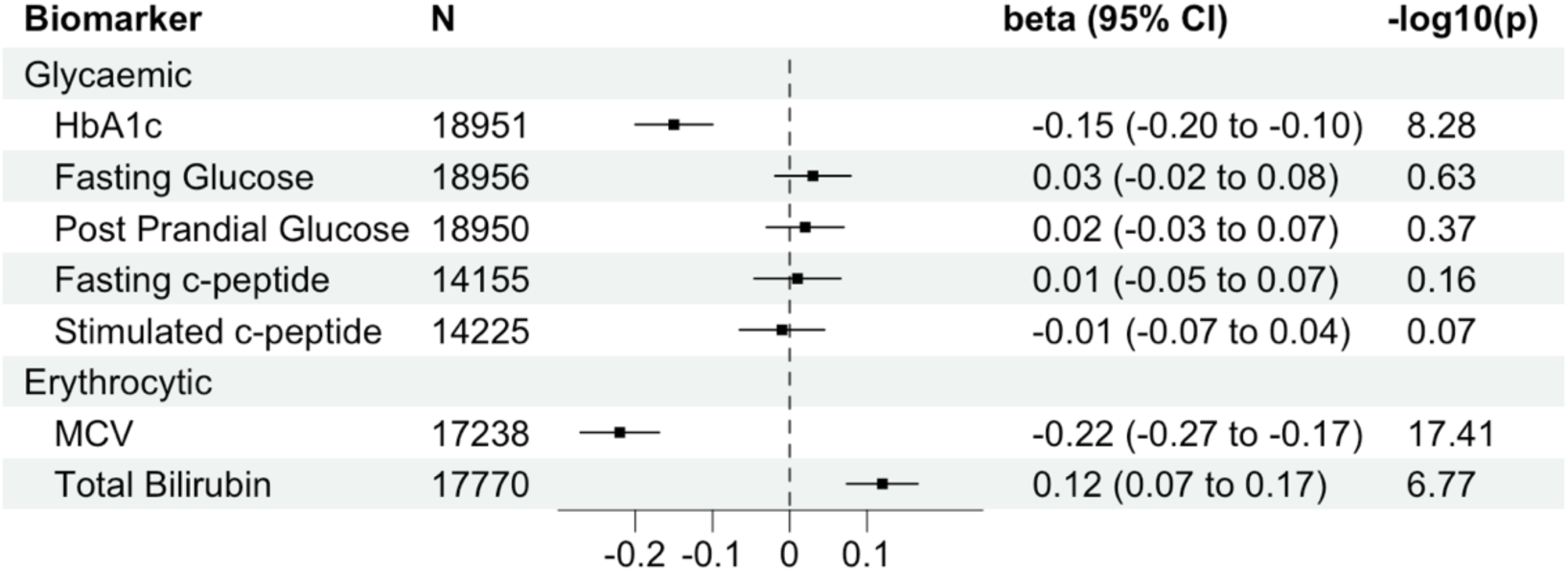
Associations between the effect allele (rs563555492T) and routinely collected biomarkers amongst South Indian participants in MDRF. Beta represents the change in standard deviation units of rank inverse normal transformed biomarkers per allele. Estimates were generated in additive genetic models adjusted for age, sex and five principal components. MCV: Mean Corpuscular Volume.

We assessed the association between rs563555492_T_ and clinical outcomes including prediabetes, T2D and diabetic eye disease using longitudinal health record date in G&H. Details of study population selection for survival analyses are available in the supplementary material (S2).

Among 31,112 individuals without prediabetes or T2D at baseline (median age 34·1 years [IQR 27·7-41·5]; median follow-up 10·9 years [10·3-11·5]), ∼8,398 (26·9%) developed prediabetes (Table 2). Incidence was 7,920 (27·7%) of 28,582 non-carrier (GG) individuals, 469 (19·1%) of 2,455 heterozygote (GT) individuals, and fewer than ten (∼12%) of 77 homozygous (TT) individuals. Median age at diagnosis increased across genotype groups (44·8 years [38·9-52·6] in GG; 46·2 [40·2-53·4] in GT; 47·1 [41·9-62·7] in TT). In flexible parametric survival models adjusted for age, sex, and genetically defined ancestry, each copy of the effect allele was associated with reduced hazard of prediabetes (HR 0·63, 95% CI 0·58-0·69; p=1·9×10⁻²⁴).

**Table 2.**
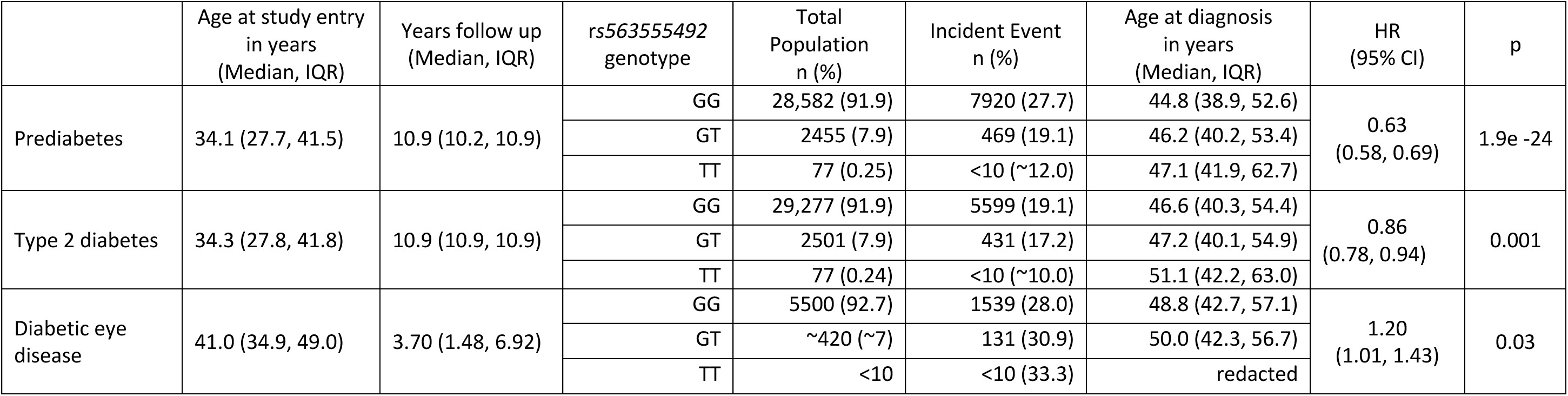
Associations between the effect allele (rs563555492T) and type 2 diabetes related outcomes from flexible parametric survival models with restricted cubic splines adjusted for age, sex and principal component-defined ancestry. Study entry for prediabetes and type 2 diabetes analyses were set as Jan 1^st^, 2014. Study entry for diabetic eye disease analysis was set as date of type 2 diabetes diagnosis. Cells containing small numbers have been rounded (∼) or redacted to reduce the chance of deidentification.

|  | Age at study entry<br>in years<br>(Median, IQR) | Years follow up<br>(Median, IQR) | *rs563555492*<br>genotype | Total<br>Population<br>n (%) | Incident Event<br>n (%) | Age at diagnosis<br>in years<br>(Median, IQR) | HR<br>(95% CI) | p |
| --- | --- | --- | --- | --- | --- | --- | --- | --- |
| Prediabetes | 34.1 (27.7, 41.5) | 10.9 (10.2, 10.9) | GG | 28,582 (91.9) | 7920 (27.7) | 44.8 (38.9, 52.6) | 0.63<br>(0.58, 0.69) | 1.9e -24 |
|  |  |  | GT | 2455 (7.9) | 469 (19.1) | 46.2 (40.2, 53.4) |  |  |
|  |  |  | TT | 77 (0.25) | <10 (~12.0) | 47.1 (41.9, 62.7) |  |  |
| Type 2 diabetes | 34.3 (27.8, 41.8) | 10.9 (10.9, 10.9) | GG | 29,277 (91.9) | 5599 (19.1) | 46.6 (40.3, 54.4) | 0.86<br>(0.78, 0.94) | 0.001 |
|  |  |  | GT | 2501 (7.9) | 431 (17.2) | 47.2 (40.1, 54.9) |  |  |
|  |  |  | TT | 77 (0.24) | <10 (~10.0) | 51.1 (42.2, 63.0) |  |  |
| Diabetic eye<br>disease | 41.0 (34.9, 49.0) | 3.70 (1.48, 6.92) | GG | 5500 (92.7) | 1539 (28.0) | 48.8 (42.7, 57.1) | 1.20<br>(1.01, 1.43) | 0.03 |
|  |  |  | GT | ~420 (~7) | 131 (30.9) | 50.0 (42.3, 56.7) |  |  |
|  |  |  | TT | <10 | <10 (33.3) | redacted |  |  |

Among 31,853 individuals without T2D at baseline (median age 34·3 years [27·8-41·8]; median follow-up 10·9 years), ∼6,040 (19·0%) developed type 2 diabetes (Table 2). Diagnosis occurred in 5,599 (19·1%) of 29,277 GG individuals, 431 (17·2%) of 2,501 GT individuals, and fewer than ten (∼10%) of 77 TT individuals. Median age at diagnosis increased across genotype groups (46·6 years [40·3-54·4] in GG, 47·2 [40·1-54·9] in GT, and 51·1 [42·2-63·0] in TT). The adjusted hazard ratio for T2D per effect allele was 0·86 (95% CI 0·78-0·94; p=0·001).

Diabetic eye disease was defined *a priori* as the principal microvascular outcome as it is an early microvascular complication, routinely screened for in clinical practice. Among ∼5,930 individuals with incident T2D (median follow-up 3·7 years [1·5-6·9]), 1,539 (28·0%) of 5,500 GG individuals developed diabetic eye disease compared with 131 (30·9%) of approximately 420 GT individuals and fewer than ten (∼33%) TT individuals (Table 2). Age at diagnosis of eye disease did not differ across genotype groups (Table 2). In adjusted survival models, each copy of the effect allele was associated with increased hazard of diabetic eye disease (HR 1·20, 95% CI 1·01-1·43; p=0·03).

We estimated 10-year absolute risks using model-derived marginal estimates in a simulated population aged 40-50 years at study entry (Figure 2, S6 Supplementary). Reduced hazards seen in variant carriers were interpreted as reflecting delayed or missed diagnosis due to non-glycaemic lowering of HbA1c. The 10-year attributable risk of missed prediabetes was 12·5% (95% CI 7·4-17·7) in GT and 21·6% (16·2-26·7) in TT individuals. Applying genotype frequencies derived from a T-allele frequency of 4·1% (7·86% GT; 0·17% TT), this corresponds to approximately 1,019 excess missed prediabetes diagnoses per 100,000 adults over 10 years (≈983 from GT; ≈36 from TT). For type 2 diabetes, the attributable risk of missed diagnosis was 3·7% (1·4-5·8) in GT and 7·1% (2·3-10·4) in TT individuals, equating to approximately 303 excess missed diagnoses per 100,000 adults over 10 years (≈291 from GT; ≈12 from TT). Amongst individuals with T2D, attributable risk of diabetic eye disease within 10 years of T2D diagnosis were 6·3% (0·5-12·2) in GT and 12·8% (0·4-25·3) in TT individuals. Applying genotype frequencies among individuals with T2D yielded approximately 517 excess cases of diabetic eye disease per 100,000 people with T2D over 10 years (≈495 from GT; ≈22 from TT), although estimates were imprecise for TT owing to small numbers (S6 Supplementary).

**Figure 2.**
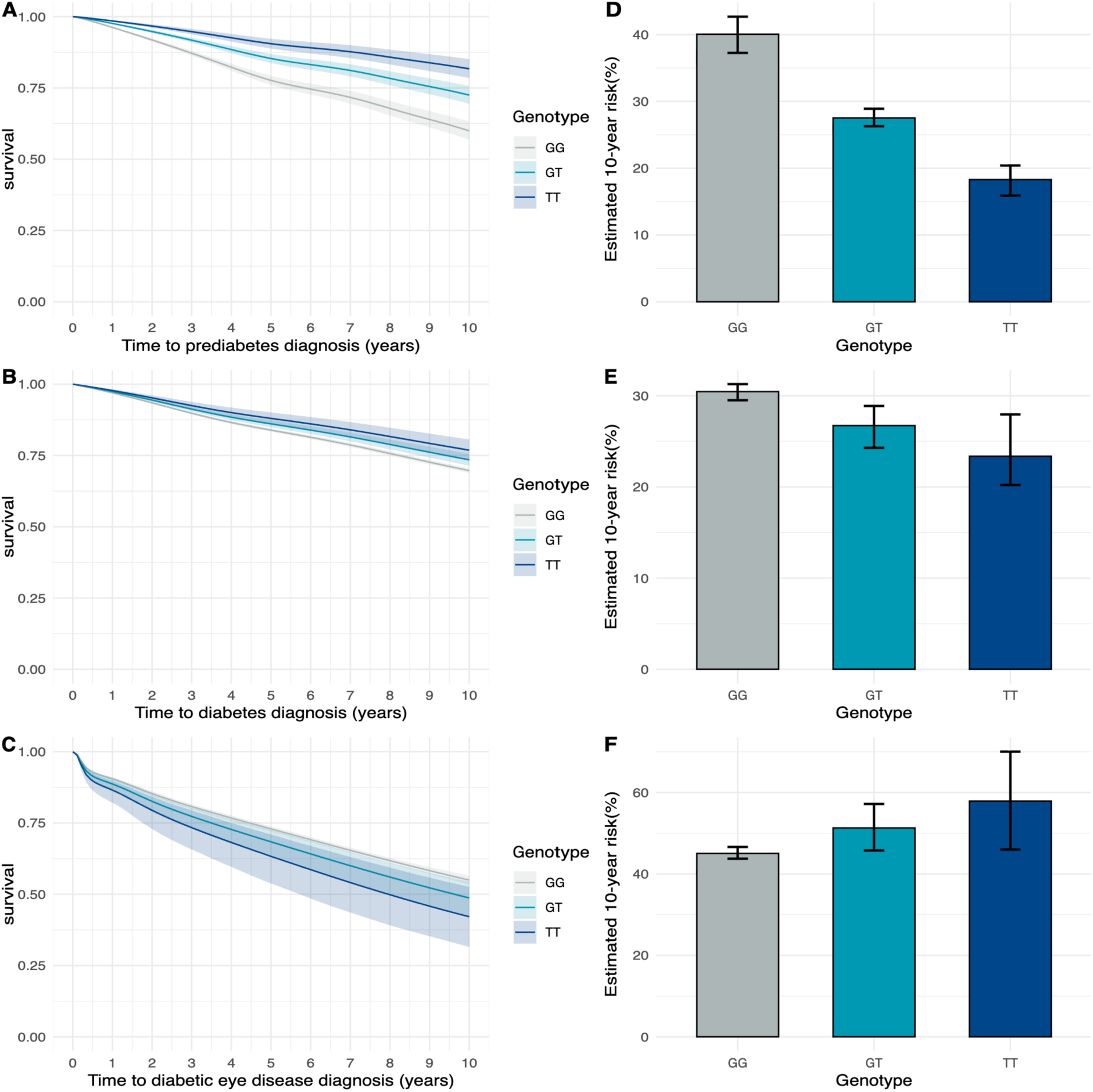
Panels A to C demonstrate marginal survival curves by effect allele (rs563555492T) status generated for a modelled population with the characteristics of the study cohort and with age set to 40-50 years at study entry on Jan 1^st^, 2014. Panels (A) and(B) show time to prediabetes and type 2 diabetes diagnosis, respectively. Panel (C) shows time from type 2 diabetes diagnosis to diabetic eye disease. Estimates from these models were used to calculate the predicted 10-year absolute risk of (D) prediabetes, (E) type 2 diabetes, and (F) diabetic eye disease.

Sensitivity analyses yielded consistent effect estimates. Stratified analyses by sex and ancestry group demonstrated consistent directions of effect. Adjustment for HbA1c in the diabetic eye disease models did not materially alter associations, although precision was reduced due to smaller sample size (S3 Supplementary).

We estimated the potential health economic impact of the 1,019 missed prediabetes diagnoses per 100,000 individuals attributable to the *rs563555492T* variant, as missed opportunities for referral into the NHS Diabetes Prevention Programme (NHS-DPP). Using published estimates of incremental QALY gains and cost savings associated with programme referral, these missed prediabetes diagnoses correspond to missed opportunities to gain approximately 41·6 QALYs and £138 336 in direct healthcare savings. Applying standard NICE cost-effectiveness thresholds (£20,000 to £30,000 per QALY) the total economic burden, comprising both direct healthcare expenditures and monetised health loss, ranges from £970,000 to £1,390,000 per 100,000 individuals over a 10-year horizon. Under these assumptions, alternative diagnostic strategies or targeted genetic screening would remain cost-effective if implementation costs were below £9·70 - £13·90 per person. For context, this is broadly comparable to (or lower than) the cost of many routine NHS blood tests.^33^

## Discussion

In this integrated analysis of genetic and health data from individuals of south Asian ancestry in the UK and India, we show that the *PIEZO1* rs563555492_T_ variant is associated with lower HbA1c independent of glycaemia, delayed diagnosis of prediabetes and type 2 diabetes, and increased risk of diabetic eye disease among individuals with type 2 diabetes. These findings provide important replication of this previously reported association in a geographically distinct South Indian cohort, and using deeper metabolic phenotyping, it builds additional evidence for the variant exerting its effect via non-glycaemic mechanisms. By harnessing longitudinal health data from south Asians in the UK, we then show that the variant is associated with missed pre-diabetes and T2D diagnosis. Based on allele frequencies, we estimate that missed diagnosis could impact one in twelve south Asian people and result in increased risk of microvascular diabetes complications and greater economic cost.

Our findings support a non-glycaemic effect on HbA1c mediated through erythrocyte biology. *PIEZO1* encodes a mechanosensitive ion channel expressed in multiple tissues. ^19,22,34^ Gain-of-function mutations in PIEZO1 cause dehydration hereditary stomatocytosis,^20^ a haemolytic disorder in which reduced HbA1c has been described in case reports.^35,36^ Such mutations have also been associated with partial resistance to malaria ^37^ suggesting evolutionary selection in malaria-endemic regions may have shaped variation in this pathway. Accordingly, the variant was associated with lower HbA1c, but not with fasting or postprandial glucose. Although animal models suggest PIEZO1 channels may influence pancreatic insulin secretion,^22^ we observed no evidence of association between the variant and C-peptide concentration, a marker of insulin secretion.^38^ These convergent findings support an erythrocytic mechanism linking rs563555492_T_ to reduced HbA1c.^13,16,17^

These findings add to the evidence that ancestry-enriched genetic determinants of erythrocyte biology can systematically bias HbA1c-based diagnosis of type 2 diabetes ^13,17,39^ and may have adverse clinical consequences.^13,14,40^ Carriers of rs563555492_T_ experienced delayed diagnosis of both prediabetes and T2D. Many health systems now aim to identify prediabetes to deliver structured diabetes prevention programmes deemed cost-effective.^5-7,23^ Our findings therefore suggest that carriers will experience delays in access to risk-modifying preventative interventions. The increased risk of diabetic eye disease observed among variant carriers with type 2 diabetes is consistent with the possibility that HbA1c underestimates glycaemia, delaying diagnosis and treatment. Increased risk of retinopathy has also been reported in association with other variants which reduce HbA1c through erythrocytic mechanisms.^14,40^

The direction of effect of rs563555492_T_ on HbA1c is concordant with that reported for other erythrocytic variants^14,40^ but contrasts with epidemiological observations that HbA1c may overestimate glycaemia in south Asian populations.^41,42^ Together, these findings underscore the importance of interpreting biomarkers in the context of individual ancestral and biological variation, rather than assuming predictable association with broad racial or ethnic categories.

We estimated the projected economic impact of delayed prediabetes detection from a UK NHS perspective. Modelling missed prediabetes diagnoses as foregone opportunities for referral to structured prevention programmes, suggested population-level QALY losses and associated net monetary impact. These projections rely on assumptions derived from published programme evaluations and do not include wider societal costs, such as productivity loss or social care needs, and should therefore be interpreted cautiously. Nevertheless, they demonstrate how even modest genotype-associated shifts in HbA1c may translate into measurable health system consequences when applied at population scale.

This study has several strengths. We integrated data from independent cohorts in South Asia and the UK, enabling replication and longitudinal evaluation across a diverse south Asian ancestral background^43^ in both migrant and non-migrant communities. The G&H and MDRF cohorts provide complementary study designs that have allowed us to build a deeper understanding of both the mechanism and clinical impact of this *PIEZO1* variant. Triangulation across genetic, and biomarker data further strengthens mechanistic inference.

This study has some limitations. The number of homozygous carriers was small, limiting precision of estimates, particularly for less common outcomes such as diabetic eye disease. Outcomes were defined using routinely collected primary care data and may be subject to misclassification, although this is unlikely to differ across randomly allocated genotypes. Economic estimates rely on assumptions regarding programme uptake and long-term effectiveness. Finally, the findings may not generalise to healthcare systems with different screening, diagnostic and intervention pathways.

In summary, a *PIEZO1* variant enriched in south Asian populations lowers HbA1c independent of glycaemia, delays clinical recognition of prediabetes and type 2 diabetes, and is associated with increased risk of diabetic eye disease. In the global south Asian populations where this variant is enriched, evaluation of complementary glycaemic biomarkers or genotype-informed HbA1c interpretation may help ensure precise and timely intervention and reduce the burden and cost of type 2 diabetes complications.

## Data Availability

Individual-level participant data are available to researchers via application to Genes & Health and MDRF

## Author Contribution

MS, DS, BJ, SM, CAR, JS, RM, TJM, MKS, IB and SF contributed to conceptualisation and methodology. MS, DS, VB, MB, JCZ and VLE contributed to formal analysis. SR, DAvH, VB, VR and JS contributed to data curation. DS, TB, TJM, VLE, MKS, IB and SF contributed to supervision. VM, RMA, DAvH, SF contributed to provision of resources. MS, RM, IB, SF contributed to funding acquisition. MS wrote the original draft. All authors reviewed and edited the draft.

## Acknowledgements

For the purpose of open access, the author has applied a Creative Commons Attribution (CC BY) licence to any Author Accepted Manuscript version arising from this submission

Genes & Health is/has recently been core-funded by Wellcome (WT102627, WT210561), the Medical Research Council (UK) (M009017, MR/X009777/1, MR/X009920/1), Higher Education Funding Council for England Catalyst, Barts Charity (845/1796), Health Data Research UK (for London substantive site), and research delivery support from the NHS National Institute for Health Research Clinical Research Network (North Thames). Genes & Health is/has recently been funded by Alnylam Pharmaceuticals, Genomics PLC; and a Life Sciences Industry Consortium of AstraZeneca PLC, Bristol-Myers Squibb Company, GlaxoSmithKline Research and Development Limited, Maze Therapeutics Inc, Merck Sharp & Dohme LLC, Novo Nordisk A/S, Pfizer Inc, Takeda Development Centre Americas Inc.

We thank Social Action for Health, Centre of The Cell, members of our Community Advisory Group, and staff who have recruited and collected data from volunteers. We thank the NIHR National Biosample Centre (UK Biocentre), the Social Genetic & Developmental Psychiatry Centre (King’s College London), Wellcome Sanger Institute, and Broad Institute for sample processing, genotyping, sequencing and variant annotation.

This work uses data provided by patients and collected by the NHS as part of their care and support. This research utilised Queen Mary University of London’s Apocrita HPC facility, supported by QMUL Research-IT, http://doi.org/10.5281/zenodo.438045

We thank: Barts Health NHS Trust, NHS Clinical Commissioning Groups (City and Hackney, Waltham Forest, Tower Hamlets, Newham, Redbridge, Havering, Barking and Dagenham), East London NHS Foundation Trust, Bradford Teaching Hospitals NHS Foundation Trust, Public Health England (especially David Wyllie), Discovery Data Service/Endeavour Health Charitable Trust (especially David Stables), Voror Health Technologies Ltd (especially Sophie Don), NHS England (for what was NHS Digital) - for GDPR-compliant data sharing backed by individual written informed consent.

A favourable ethical opinion for the main Genes & Health research study was granted by NRES Committee London - South East (reference 14/LO/1240) on 16 Sept 2014. Queen Mary University of London is the Sponsor.

This study was supported by the National Institute for Health and Care Research Exeter Biomedical Research Centre. The views expressed are those of the authors and not necessarily those of the NIHR or the Department of Health and Social Care.

We acknowledge the support of Indian Council of Medical Research (ICMR) for setting up the MDRF - Biobank

Most of all we thank all the volunteers participating in MDRF and Genes & Health.

## S1. Electronic Health Record (EHR) data curation in Genes & Health

Clinical outcomes were defined in Genes & Health using curated codelists for prediabetes, type 2 diabetes and diabetic eye disease that have previously been used and validated.^1^ We excluded individuals with clinical codes suggestive of type 1 diabetes, monogenic diabetes, MODY, and diabetes secondary to cystic fibrosis or pancreatic disease.

**S1 Table.**
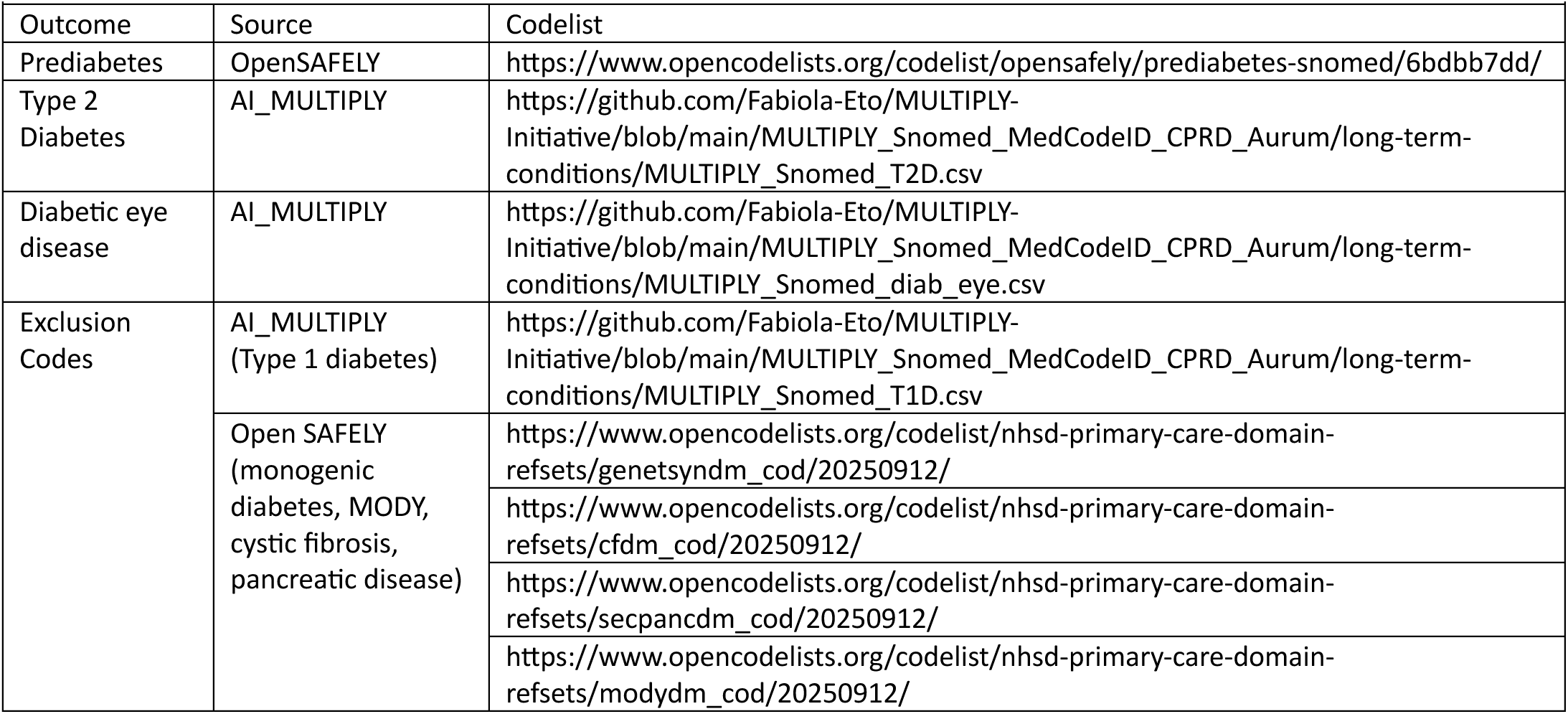

| S1 Table |  |  |
| --- | --- | --- |
| Outcome | Source | Codelist |
| Prediabetes | OpenSAFELY | https://www.opencodelists.org/codelist/opensafely/prediabetes-snomed/6bdbb7dd/ |
| Type 2 Diabetes | AI_MULTIPLY | https://github.com/Fabiola-Eto/MULTIPLY-Initiative/blob/main/MULTIPLY_Snomed_MedCodeID_CPRD_Aurum/long-term-conditions/MULTIPLY_Snomed_T2D.csv |
| Diabetic eye disease | AI_MULTIPLY | https://github.com/Fabiola-Eto/MULTIPLY-Initiative/blob/main/MULTIPLY_Snomed_MedCodeID_CPRD_Aurum/long-term-conditions/MULTIPLY_Snomed_diab_eye.csv |
| Exclusion Codes | AI_MULTIPLY (Type 1 diabetes) | https://github.com/Fabiola-Eto/MULTIPLY-Initiative/blob/main/MULTIPLY_Snomed_MedCodeID_CPRD_Aurum/long-term-conditions/MULTIPLY_Snomed_T1D.csv |
|  | Open SAFELY (monogenic diabetes, MODY, cystic fibrosis, pancreatic disease) | https://www.opencodelists.org/codelist/nhsd-primary-care-domain-refsets/genetsyndm_cod/20250912/ |
|  |  | https://www.opencodelists.org/codelist/nhsd-primary-care-domain-refsets/cfdm_cod/20250912/ |
|  |  | https://www.opencodelists.org/codelist/nhsd-primary-care-domain-refsets/secpancdm_cod/20250912/ |
|  |  | https://www.opencodelists.org/codelist/nhsd-primary-care-domain-refsets/modydm_cod/20250912/ |

Data from the National Diabetes Audit (NDA)^2^, an annual England-wide registry of patients with diabetes, were used to refine date of diagnosis data and exclusion criteria. If there was a mismatch between the first recorded code available in Genes & Health and that in the NDA, the NDA date was taken preferentially. Additionally, if individuals had a diabetes type that was not consistent with type 2 diabetes, they were excluded.

Codelists used to define further diabetes complications are available via the AI_MULTIPLY initiative^3^.

## S2. Study population selection for survival analyses

Selection of the study sample for each stage of the survival analysis in Genes & Health. Numbers included and excluded in diabetic eye disease analysis have been rounded to prevent subsequent low cell count disclosure.

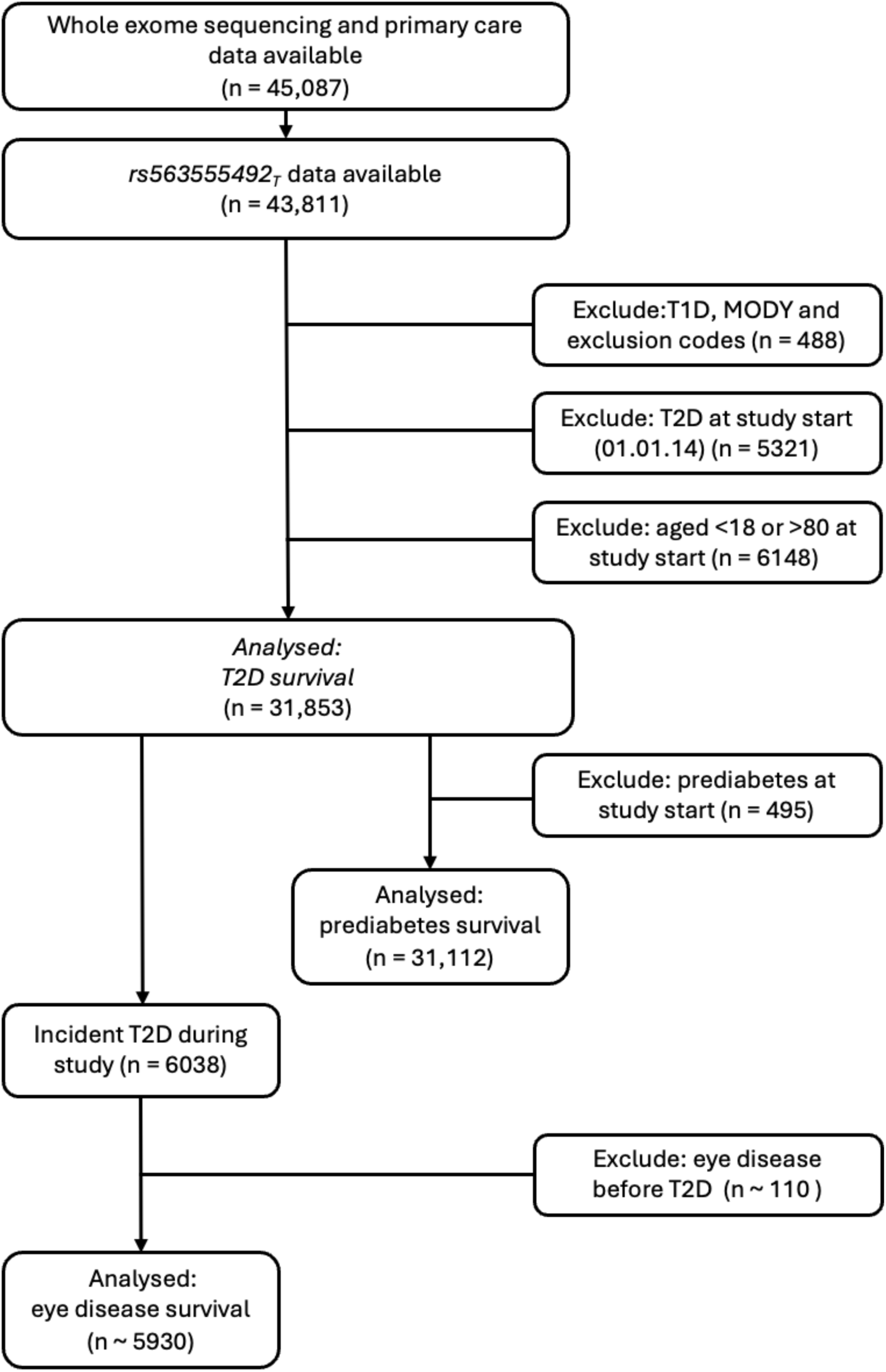

## S3. Survival Model Development

### Study design and start of follow-up

HbA1c has been used for monitoring type 2 diabetes since assay standardisation in the 1990s and for diagnosis since the 2011 WHO recommendation. Because our primary hypothesis concerned the association between the rs563555492_T_ and study outcomes through its effect on HbA1c we selected a start date after these recommendations were published and following implementation of the NHS Health Check programme, a national cardiometabolic screening intervention introduced over a 5-year period from 2009. Follow-up for survival analyses therefore commenced on Jan 1, 2014. Diabetic eye disease was defined *a priori* as the primary microvascular outcome as it is a common outcome that is routinely screened for in an NHS setting.

### Statistical analysis

We first examined unadjusted associations using Kaplan–Meier curves.

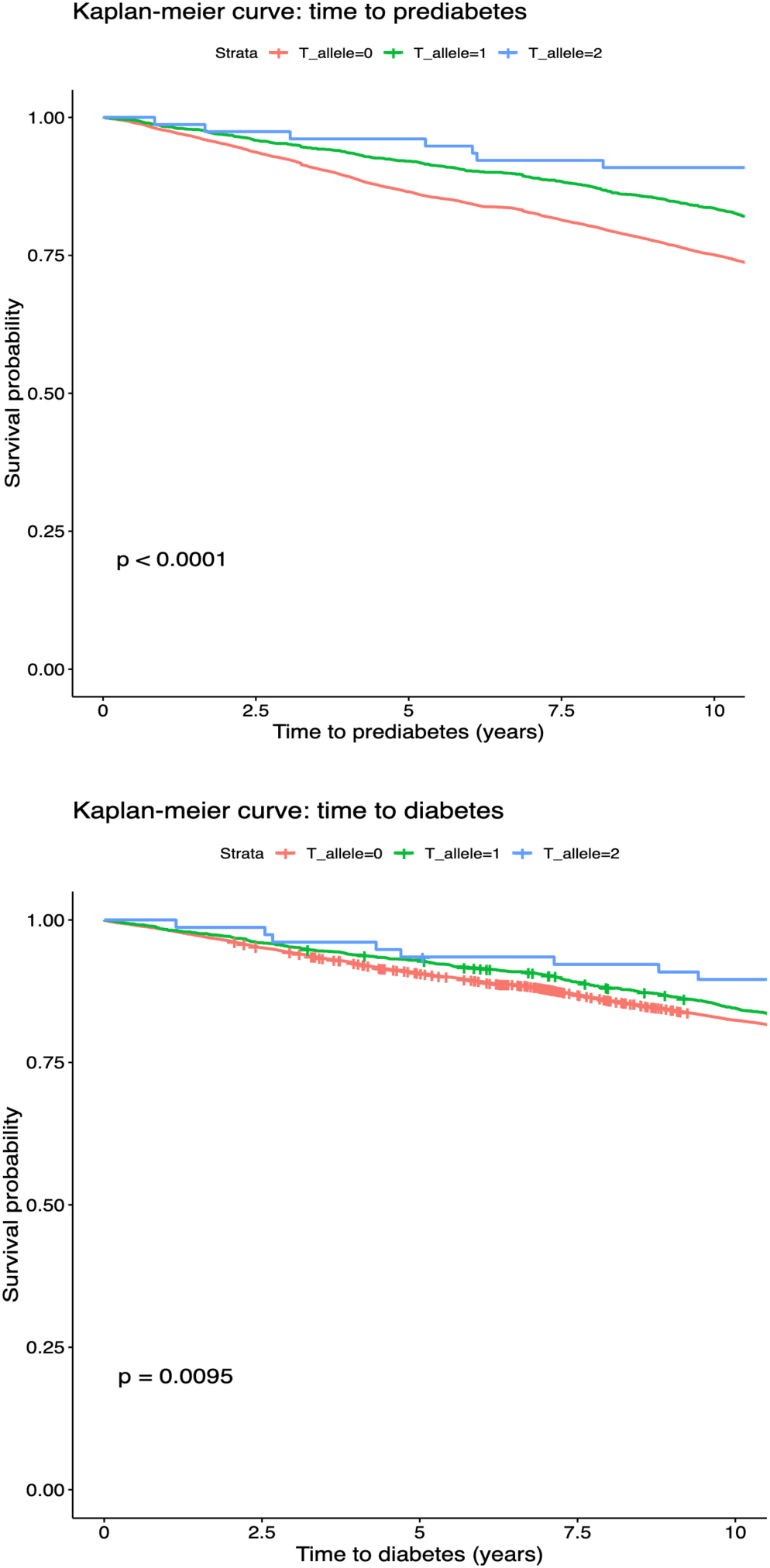

Individuals with the TT genotype have been redacted from the figure below to reduce risk of disclosure.

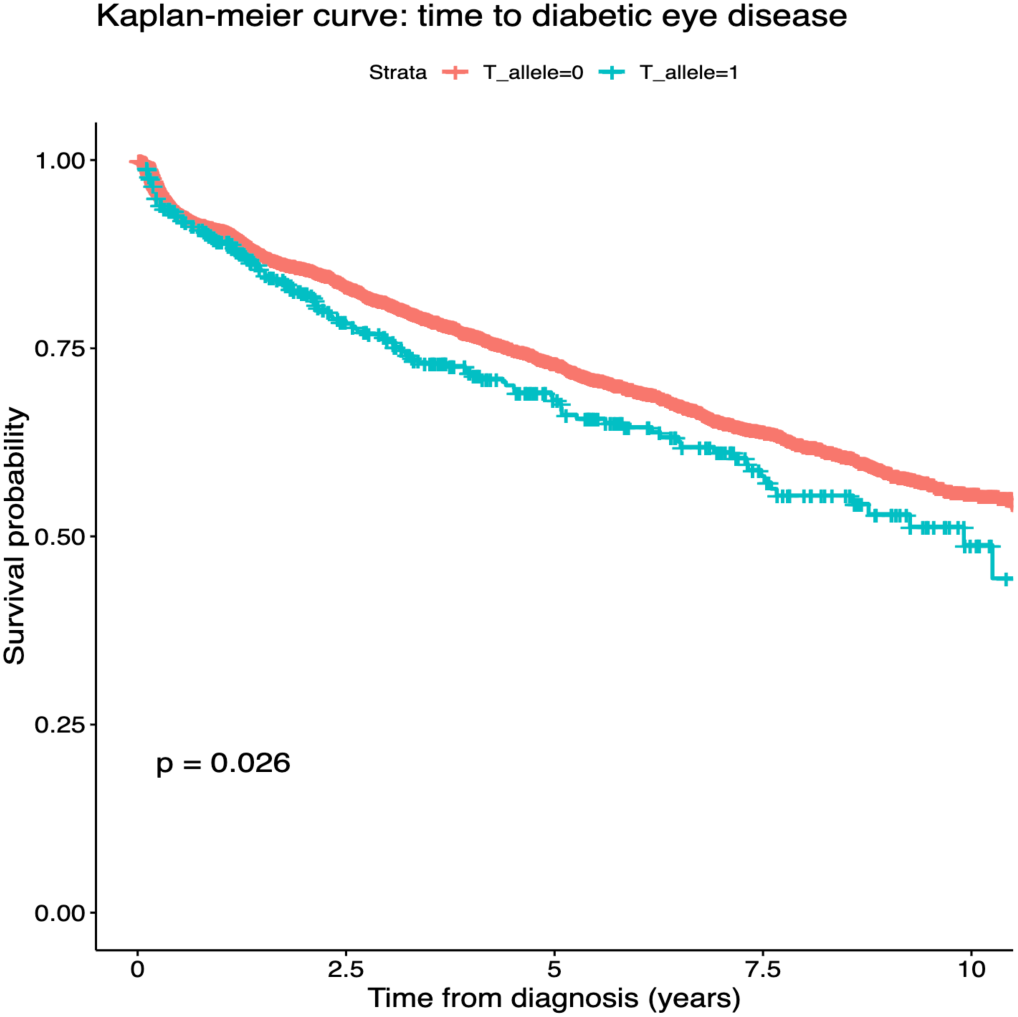

Associations between rs563555492_T_ and each outcome were then assessed using Cox proportional hazards models. Covariates considered potential confounders of the association between rs563555492_T_ and type 2 diabetes–related outcomes were specified a priori. Because age demonstrated a non-linear association with each outcome, restricted cubic spline terms were included in multivariable models. We compared two adjustment strategies. Model 1 included age, sex, and 20 genetic principal components (PCs) used in the discovery analysis. Model 2 included age, sex, and binarised PC-derived ancestry (Bangladeshi or Pakistani). As effect estimates were similar between models (Supplementary Table 2), we present results from Model 2 in the main analyses to maximise statistical power in outcomes with small event numbers.

**S3a Table.** Comparison of Cox models used to generate estimate effect of rs563555492_T_ on longitudinal outcomes. Model 1 and 2 are adjusted for sex and age at study entry. Model 1 is adjusted for 20 genetic principal components. Model 2 is adjusted for a composite exposure ‘ancestry’ of principal components derived from ancestry. A spline term for age has been used in both Model 1 and Model 2 to account for variation in the estimated effect of age on the outcome across ages.

|  | Prediabetes |  | Type 2 Diabetes |  | Eye |  |
| --- | --- | --- | --- | --- | --- | --- |
|  | HR (95% CI) | p | HR (95% CI) | p | HR (95% CI) | p |
| Model 1 | 0.63 (0.58, 0.69) | 3.5e -23 | 0.86 (0.78, 0.94) | 1.94e -3 | 1.19 (0.998, 1.41) | 0.053 |
| Model 2 | 0.63 (0.57, 0.69) | 1.01e -24 | 0.85 (0.78, 0.94) | 9.75e -4 | 1.20 (1.01, 1.43) | 0.036 |

### Testing of proportional hazards assumption

We tested proportional hazards assumptions for each Cox model using the cox.zph command from the R survival package. There was no meaningful evidence that the rs563555492 genotype violated the proportional hazards assumption; however, some covariates demonstrated evidence of non- proportionality, suggesting that their effect on the outcome varied over time (Supplementary Table 3).

**S3b Table.**
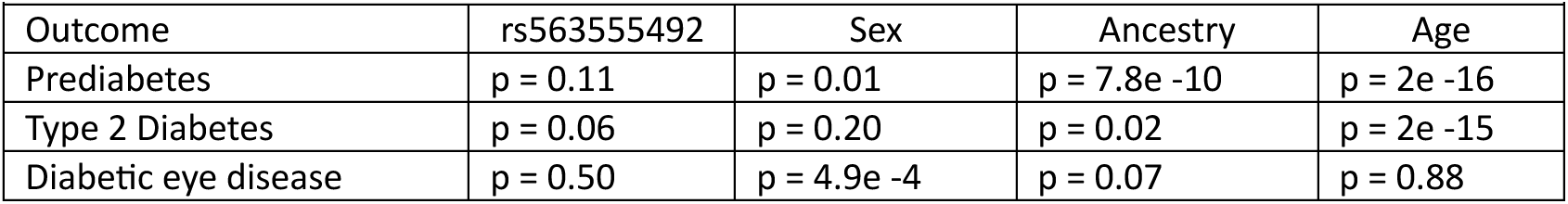
Evidence that specific covariates violate the proportional hazards assumption for each outcome. P values were generated using cox.zph.

| Outcome | rs563555492 | Sex | Ancestry | Age |
| --- | --- | --- | --- | --- |
| Prediabetes | p = 0.11 | p = 0.01 | p = 7.8e -10 | p = 2e -16 |
| Type 2 Diabetes | p = 0.06 | p = 0.20 | p = 0.02 | p = 2e -15 |
| Diabetic eye disease | p = 0.50 | p = 4.9e -4 | p = 0.07 | p = 0.88 |

### Stratified analyses

Sex and ancestry were strongly associated with study outcomes, and each variable violated proportional hazards assumptions in some models. We therefore conducted sensitivity analyses stratified by sex and binarised PC-derived ancestry. Although analyses for diabetic eye disease were underpowered in some subgroups, particularly among participants of Pakistani ancestry, effect estimates were directionally consistent across strata.

**S3c Table.** Estimated effect of rs563555492_T_ on risk of study outcomes in ancestry stratified analysis

|  |  |  |  |  |  |  |
| --- | --- | --- | --- | --- | --- | --- |
|  | Prediabetes |  | Diabetes |  | Eye Disease |  |
|  | Hazard Ratio (95% CI) | p value | Hazard Ratio (95% CI) | p value | Hazard Ratio (95% CI) | p value |
| Bangladeshi | 0.65 (0.59, 0.73) | 2.2e-15 | 0.87(0.78, 0.97) | 0.012 | 1.24 (1.02, 1.52) | 0.029 |
| Pakistani | 0.57 (0.48, 0.68) | 1.4e-10 | 0.80 (0.66, 0.97) | 0.021 | 1.08 (0.73, 1.57) | 0.7 |

### Sensitivity Analysis

To explore whether associations were mediated through glycaemia, we additionally adjusted Cox models for the most recent HbA1c measured on or before the recorded date of type 2 diabetes diagnosis. Models were also adjusted for age, sex, and binarised PC-derived ancestry. An additional 640 individuals were excluded due to absence of an HbA1c measurement recorded on or before diagnosis. HbA1c measures recorded in the electronic health record of Genes & Health participants were cleaned and linked to genomic data using methods described previously (Martin, Samuel et al. 2025). There was no evidence of a difference in HbA1c across genotype groups (Kruskal–Wallis χ²(2) = 0·86, P = 0·65). Adjustment for HbA1c attenuated statistical power but did not alter the direction of effect of rs563555492_T_ on diabetic eye disease risk (Supplementary Table 5).

### Development of flexible parametric models

We have used flexible parametric survival models with restricted cubic splines to allow for variation in time varying effects within the splines and to allow post model estimations of risk of outcomes. Model development proceeded as follows: (1) the baseline hazard was modelled for each outcome; (2) Model fit statistics (log-likelihood, AIC, and BIC) were compared across models incorporating 0–10 knots to determine the optimal specification; and (3) for covariates demonstrating non-proportional hazards, time- varying effects were incorporated within spline terms. Estimates from flexible parametric models were consistent with those generated in the cox models for each outcome.

**S3d Table.** Sensitivity analysis exploring association between genotype and risk of diabetic eye disease in a model adjusted for HbA1c at the time of diagnosis

|  |  |  |  |  |  |
| --- | --- | --- | --- | --- | --- |
|  |  | HbA1c (mmol/mol) |  | Diabetic Eye Disease |  |
| rs563555492 Genotype | N | Median (IQR) | P value (Kruskal-Wallis) | Hazards Ratio (95% CI) | p value |
| GG | 5500 | 51.0 (48.0, 58.0) | 0.65 | 1.10 (0.95, 1.40) | 0.14 |
| GT | 424 | 52.0 (48.0, 60.0) |  |  |  |
| TT | <10 | redacted |  |  |  |

## S4. Associations between rs563555492 and type 2 diabetes related outcomes

**S4 Table.** Estimates generated in flexible parametric models with restricted cubic splines adjusted for age, sex and a binarized principal component-based ancestry. Age was modelled using restricted cubic splines to allow for non-linearity, therefore estimates for the effect of age on the outcome are not presented.

|  | Prediabetes |  |  |  | Type 2 diabetes |  |  |  | Diabetic eye disease |  |  |  |
| --- | --- | --- | --- | --- | --- | --- | --- | --- | --- | --- | --- | --- |
|  | Population | Events |  |  | Population | Events |  |  | Population | Events |  |  |
|  | N (%) | n (%) | HR (95%CI) | p | N (%) | n (%) | HR (95% CI) | p | N (%) | n (%) | HR (95% CI) | p |
| **Genotype** |  |  |  |  |  |  |  |  |  |  |  |  |
| GG | 28,582 (91.9) | 7920 (27.7) | 0.63<br>(0.58, 0.69) | 1.94e -24 | 29,277 (91.9) | 5599 (19.1) | 0.86<br>(0.78, 0.94) | 0.001 | 5500 (92.7) | 1539 (28.0) | 1.20<br>(1.01, 1.43) | 0.03 |
| GT | 2455 (7.9) | 469 (19.1) |  |  | 2501 (7.9) | 431 (17.2) |  |  | ~420 (~7) | 131 (30.9) |  |  |
| TT | 77 (0.25) | <10 (~12.0) |  |  | 77 (0.24) | <10 (~10.0) |  |  | <10 | <10 (33.3) |  |  |
| **Sex** |  |  |  |  |  |  |  |  |  |  |  |  |
| Female | 17,158 (55.1) | 4460 (26.0) | ref |  | 17,591(55.2) | 2854 (16.2) | ref |  | 2798 (47.2) | 685 (24.5) | ref |  |
| Male | 13,956 (44.9) | 3938 (28.2) | 0.93<br>(0.88, 0.97) | 3.3e -4 | 14,264 (44.8) | 3184 (22.3) | 1.14<br>(1.08, 1.20) | 4.86e -7 | 3132 (52.8) | 987 (31.5) | 1.30<br>(1.18, 1.43) | 2.0 e -7 |
| **Ancestry** |  |  |  |  |  |  |  |  |  |  |  |  |
| Bangladeshi | 20,627 (66.3) | 5680 (27.5) | ref |  | 21,224 (66.6) | 4394 (20.7) |  |  | 4317 (72.8) | 1272 (29.5) |  |  |
| Pakistani | 10,487 (33.7) | 2718 (25.9) | 0.80<br>(0.76, 0.84) | 2.2e -24 | 10,631 (33.4) | 1644 (15.5) | 0.63<br>(0.59, 0.66) | 5.46e -56 | 1613 (27.2) | 400 (24.8) | 0.78<br>(0.69, 0.87) | 2.0 e -5 |

## S5. Frequency of type 2 diabetes complications

**S5 Table.** Frequency of complications amongst individuals diagnosed with type 2 diabetes after the study start date (1^st^ January 2014), without the specified complication on or before the date of type 2 diabetes diagnosis, stratified by r*s563555492* genotype. The *combined microvascular* complication includes chronic kidney disease, neuropathy and diabetic eye disease. The *combined macrovascular* complication includes chronic heart disease, stroke, peripheral arterial disease and combined microvascular disease. The initial N varies between outcomes due to variations in the number of individuals with each outcome prior to their type 2 diabetes diagnosis.

|  | Complication | rs563555492 Genotype | N | n (events) | % (events) |
| --- | --- | --- | --- | --- | --- |
| Microvascular Complications | Chronic Kidney Disease | GG | 5649 | 229 | 4.1 |
|  |  | GT | 434 | 15 | 3.5 |
|  |  | TT | <10 | <10 | redacted |
|  | Neuropathy | GG | 5870 | 96 | 1.6 |
|  |  | GT | 447 | <10 | redacted |
|  |  | TT | <10 | 0 | 0 |
|  | Combined Microvascular | GG | 5565 | 1707 | 30.7 |
|  |  | GT | 447 | 136 | 31.8 |
|  |  | TT | <10 | <5 | 33.3 |
| Macrovascular complications | Chronic Heart Disease | GG | 6031 | 706 | 11.7 |
|  |  | GT | 463 | 59 | 12.7 |
|  |  | TT | <10 | <5 | redacted |
|  | Stroke | GG | 6031 | 111 | 1.8 |
|  |  | GT | 463 | 13 | 2.8 |
|  |  | TT | <10 | 0 | 0 |
|  | Peripheral Arterial Disease | GG | 6031 | 52 | 0.9 |
|  |  | GT | 463 | <5 | redacted |
|  |  | TT | <10 | 0 | 0 |
|  | Combined Macrovascular | GG | 6031 | 811 | 13.5 |
|  |  | GT | 463 | 68 | 14.7 |
|  |  | TT | <10 | <5 | redacted |

## S6.10-year risk and attributable risk of complications

**S6 Table.**
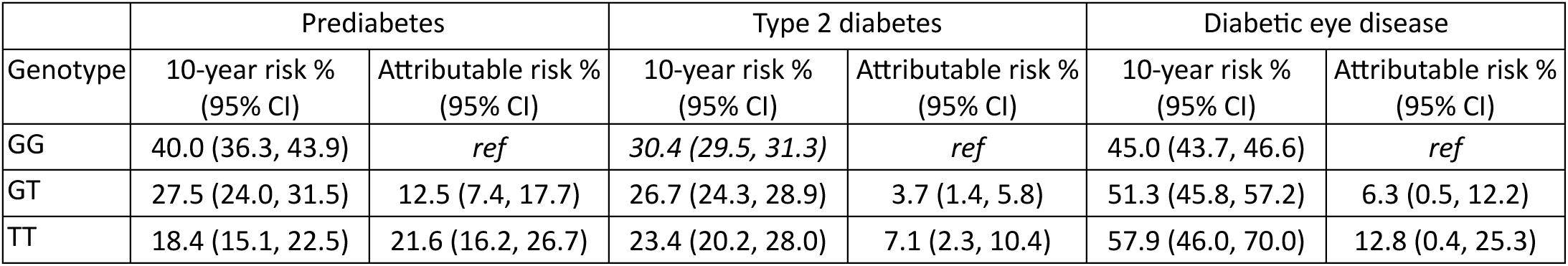
Estimated 10-year risk of outcomes generated from marginal survival curves generated for a modelled study population with age set to 40-50 years. Excess attributable risk associated with rs563555492_T_ is presented in comparison with estimates for individuals who are not carrying any copies of the effect allele (GG). For prediabetes and type 2 diabetes this is the attributable risk of missed diagnosis. For diabetic eye disease this is the excess risk of diagnosis.

## Footnotes

1 Ethnic differences in early onset multimorbidity and associations with health service use, long-term prescribing, years of life lost, and mortality: A cross-sectional study using clustering in the UK Clinical Practice Research Datalink Eto F, Samuel M, Henkin R, Mahesh M, Ahmad T, et al. (2023) Ethnic differences in early onset multimorbidity and associations with health service use, long-term prescribing, years of life lost, and mortality: A cross-sectional study using clustering in the UK Clinical Practice Research Datalink. PLOS Medicine 20(10): e1004300. https://doi.org/10.1371/journal.pmed.1004300

2 Holman, N. et al. Completion of annual diabetes care processes and mortality: A cohort study using the National Diabetes Audit for England and Wales. Diabetes Obes. Metab. 23, 2728–2740 (2021)

3 Eto, F et al. (2023). f-eto/MULTIPLY-Initiative: Version 1.1 (v1.1). Zenodo. https://doi.org/10.5281/zenodo.7643566

## References

1. Magliano DJ, Boyko EJ, IDF Diabetes Atlas 10th edition scientific committee. Chapter 3, Global picture. IDF DIABETES ATLAS. 10th ed. Brussels; 2021.

2. Hills AP, Arena R, Khunti K, et al. Epidemiology and determinants of type 2 diabetes in south Asia. The Lancet Diabetes & Endocrinology 2018; 6(12): 966–78.

3. Hodgson S, Williamson A, Bigossi M, et al. Genetic basis of early onset and progression of type 2 diabetes in South Asians. Nature Medicine 2025; 31(1): 323–31.

4. Misra A, Sattar N, Ghosh A, Nassar M, Jayawardena R, Gupta R. Type 2 diabetes in South Asians. BMJ 2025; 390: e079801.

5. McManus E. Evaluating the Long-Term Cost-Effectiveness of the English NHS Diabetes Prevention Programme using a Markov Model. Pharmacoecon Open 2024; 8(4): 569–83.

6. Vazquez Arreola E, Gong Q, Hanson RL, et al. Prediabetes remission and cardiovascular morbidity and mortality: post-hoc analyses from the Diabetes Prevention Program Outcome study and the DaQing Diabetes Prevention Outcome study. The Lancet Diabetes & Endocrinology 2026; 14(2): 137–48.

7. Echouffo-Tcheugui JB, Perreault L, Ji L, Dagogo-Jack S. Diagnosis and Management of Prediabetes: A Review. JAMA 2023; 329(14): 1206–16.

8. John WG, Hillson R, Alberti SG. Use of haemoglobin A1c (HbA1c) in the diagnosis of diabetes mellitus. The implementation of World Health Organisation (WHO) guidance 2011. Practical Diabetes 2012; 29(1): 12-a.

9. World Health Organisation. WHO Guidelines Approved by the Guidelines Review Committee. Use of Glycated Haemoglobin (HbA1c) in the Diagnosis of Diabetes Mellitus: Abbreviated Report of a WHO Consultation. Geneva: World Health Organization Copyright © World Health Organization 2011.; 2011.

10. Anjana RM, Unnikrishnan R, Deepa M, et al. Achievement of guideline recommended diabetes treatment targets and health habits in people with self-reported diabetes in India (ICMR-INDIAB-13): a national cross-sectional study. The Lancet Diabetes & Endocrinology 2022; 10(6): 430–41.

11. Valabhji J, Barron E, Bradley D, et al. Early Outcomes From the English National Health Service Diabetes Prevention Programme. Diabetes Care 2020; 43(1): 152–60.

12. Hanna FW, Wilkie V, Issa BG, Fryer AA. Revisiting screening for type 2 diabetes mellitus: the case for and against using HbA1c. British Journal of General Practice 2015; 65(633): e278–e80.

13. Jacobs BM, Stow D, Hodgson S, et al. Genetic architecture of routinely acquired blood tests in a British South Asian cohort. Nature Communications 2024; 15(1): 8929.

14. Martin S, Samuel M, Stow D, et al. Undiagnosed G6PD Deficiency in Black and Asian Individuals Is Prevalent and Contributes to Health Inequalities in Type 2 Diabetes Diagnosis and Complications. Diabetes Care 2025; 48(11): 1932–41.

15. gnomAD browser. https://gnomad.broadinstitute.org/variant/16-88716656-G-T?dataset=gnomad_r4.

16. Chen MH, Raffield LM, Mousas A, et al. Trans-ethnic and Ancestry-Specific Blood- Cell Genetics in 746,667 Individuals from 5 Global Populations. Cell 2020; 182(5): 1198–213.e14.

17. Sun Q, Graff M, Rowland B, et al. Analyses of biomarker traits in diverse UK biobank participants identify associations missed by European-centric analysis strategies. J Hum Genet 2022; 67(2): 87–93.

18. Saotome K, Murthy SE, Kefauver JM, Whitwam T, Patapoutian A, Ward AB. Structure of the mechanically activated ion channel Piezo1. Nature 2018; 554(7693): 481-6.

19. Murthy SE, Dubin AE, Patapoutian A. Piezos thrive under pressure: mechanically activated ion channels in health and disease. Nature Reviews Molecular Cell Biology 2017; 18(12): 771–83.

20. Albuisson J, Murthy SE, Bandell M, et al. Dehydrated hereditary stomatocytosis linked to gain-of-function mutations in mechanically activated PIEZO1 ion channels. Nat Commun 2013; 4: 1884.

21. Ma S, Cahalan S, LaMonte G, et al. Common PIEZO1 Allele in African Populations Causes RBC Dehydration and Attenuates Plasmodium Infection. Cell 2018; 173(2): 443–55.e12.

22. Ye Y, Barghouth M, Dou H, et al. A critical role of the mechanosensor PIEZO1 in glucose-induced insulin secretion in pancreatic β-cells. Nat Commun 2022; 13(1): 4237.

23. National Institute for Health and Care Excellence. National Institute for Health and Care Excellence. Type 2 diabetes: prevention in people at high risk.. 2017. https://www.nice.org.uk/guidance/ph382026).

24. Siddiqui MK, Anjana RM, Dawed AY, et al. Young-onset diabetes in Asian Indians is associated with lower measured and genetically determined beta cell function. Diabetologia 2022; 65(6): 973–83.

25. Srinivasan S, Liju S, Sathish N, et al. Common and Distinct Genetic Architecture of Age at Diagnosis of Diabetes in South Indian and European Populations. Diabetes Care 2023; 46(8): 1515–23.

26. Siddiqui MK, Dupuis T, Anjana RM, et al. XBP1 expression in pancreatic islet cells is associated with poor glycaemic control especially in young non-obese onset diabetes across ancestries. Communications Medicine 2025; 5(1): 396.

27. Finer S, Martin HC, Khan A, et al. Cohort Profile: East London Genes & Health (ELGH), a community-based population genomics and health study in British Bangladeshi and British Pakistani people. Int J Epidemiol 2020; 49(1): 20–1i.

28. Hodgson S, Bui V, Bigossi M, et al. Exome-wide association study in 54,698 south Asians identifies novel type 2 diabetes associations with RNF19A, HNF4A, and dissects role of coding variants in GP2 and CDKAL1. medRxiv 2025: 2025.09.24.25336527.

29. Anjana RM, Baskar V, Nair ATN, et al. Novel subgroups of type 2 diabetes and their association with microvascular outcomes in an Asian Indian population: a data-driven cluster analysis: the INSPIRED study. BMJ Open Diabetes Research & Care 2020; 8(1): e001506.

30. Mbatchou J, Barnard L, Backman J, et al. Computationally efficient whole-genome regression for quantitative and binary traits. Nature Genetics 2021; 53(7): 1097–103.

31. Robson J, Dostal I, Sheikh A, et al. The NHS Health Check in England: an evaluation of the first 4 years. BMJ Open 2016; 6(1): e008840.

32. Royston P, Parmar MK. Flexible parametric proportional-hazards and proportional- odds models for censored survival data, with application to prognostic modelling and estimation of treatment effects. Stat Med 2002; 21(15): 2175–97.

33. NHS England. National Cost Collection Data Publication. 26 Jan 2026 2024/25. https://www.england.nhs.uk/publication/2024-25-national-cost-collection-data-publication/ (accessed 28th February 2026.

34. Cahalan SM, Lukacs V, Ranade SS, Chien S, Bandell M, Patapoutian A. Piezo1 links mechanical forces to red blood cell volume. Elife 2015; 4.

35. Nakatani R, Murata T, Usui T, et al. Importance of the Average Glucose Level and Estimated Glycated Hemoglobin in a Diabetic Patient with Hereditary Hemolytic Anemia and Liver Cirrhosis. Intern Med 2018; 57(4): 537–43.

36. Song A, Lu L, Li Y, et al. Low HbA1c With Normal Hemoglobin in a Diabetes Patient Caused by PIEZO1 Gene Variant: A Case Report. Front Endocrinol (Lausanne) 2020; 11: 356.

37. Frederiksen H. Dehydrated hereditary stomatocytosis: clinical perspectives. J Blood Med 2019; 10: 183–91.

38. Leighton E, Sainsbury CA, Jones GC. A Practical Review of C-Peptide Testing in Diabetes. Diabetes Ther 2017; 8(3): 475–87.

39. Wheeler E, Leong A, Liu CT, et al. Impact of common genetic determinants of Hemoglobin A1c on type 2 diabetes risk and diagnosis in ancestrally diverse populations: A transethnic genome-wide meta-analysis. PLoS Med 2017; 14(9): e1002383.

40. Breeyear JH, Hellwege JN, Schroeder PH, et al. Adaptive selection at G6PD and disparities in diabetes complications. Nat Med 2024; 30(9): 2480–8.

41. Nazir A, Papita R, Anbalagan VP, Anjana RM, Deepa M, Mohan V. Prevalence of diabetes in Asian Indians based on glycated hemoglobin and fasting and 2-H post-load (75-g) plasma glucose (CURES-120). Diabetes Technol Ther 2012; 14(8): 665–8.

42. Eastwood SV, Tillin T, Mayet J, et al. Ethnic differences in cross-sectional associations between impaired glucose regulation, identified by oral glucose tolerance test or HbA1c values, and cardiovascular disease in a cohort of European and South Asian origin. Diabet Med 2016; 33(3): 340–7.

43. Kerdoncuff E, Skov L, Patterson N, et al. 50,000 years of evolutionary history of India: Impact on health and disease variation. Cell 2025; 188(13): 3389–404.e6.

